# Periodontal therapy on disease activity of Rheumatoid Arthritis. Systematic review and meta-analysis

**DOI:** 10.1101/2020.05.25.20111328

**Authors:** Francisco Novillo, Shuheng Lai, Geovanna Cárdenas, Francisca Verdugo, Gabriel Rada

## Abstract

**Objective:** The objective of this systematic review is to assess the impact of periodontal therapy on disease activity of patients with Rheumatoid Arthritis

**Data Sources:** We will conduct a comprehensive search in PubMed/Medline, Embase, Cochrane Central Register of Controlled Trials (CENTRAL), Lilacs, the International Clinical Trials Registry Platform (ICTRP), ClinicalTrials.gov, US National Institutes of Health (NIH) and grey literature, to identify all relevant randomized controlled trials regardless of language or publication status (published, unpublished, in press and in progress).

**Elegibility Criteria for Selecting Studies and Methods:** We will include randomised trials evaluating the effect of periodontal therapy on disease activity of rheumatoid arthritis. Two reviewers will independently screen each study for eligibility, data extraction, and assess the risk of bias. We will pool the results using meta-analysis and will apply the GRADE system to assess the certainty of the evidence for each outcome.

**Ethics and Dissemination:** No ethics approval is considered necessary. The results of this review will be disseminated via peer-reviewed publications, social networks and traditional media.

**PROSPERO Registration ID:** CRD42020150286.

## Introduction

Rheumatoid arthritis (RA) is a chronic inflammatory joint disease, which can cause cartilage and bone damage as well as disability. RA has an incidence of 0.5% to 1%(1). The clinical manifestations of joint involvement include arthralgia, swelling, redness, and even limiting the range of motion alongside radiographic joint space narrowing and bony erosions(2). These signs and symptoms can greatly alter the quality of life of RA patients on different domains such as physical health, level of independence and personal beliefs amongst others(3). The current treatment principles for established RA involve symptomatic management and disease modification with disease modifying antirheumatic drugs (DMARDs)(2).

Periodontitis is defined as an inflammatory disease of supporting tissues of teeth caused by specific microorganisms or groups of specific microorganisms, resulting in progressive destruction of the periodontal ligament and alveolar bone with periodontal pocket formation, gingival recession or both(4). Periodontal disease (PD) is estimated to affect about 20-50% of the population around the globe(5). Periodontal treatment (PT) utilizes a plethora of therapeutic interventions, including behavioural-change approaches in addition to subgingival instrumentation to remove plaque and calculus, local and systemic pharmacotherapy, and various types of surgery(6).

PD has been proposed as having an etiologic or modulating role in different diseases such as diabetes, and adverse pregnancy outcome. Several mechanisms have been proposed to explain or support such theories(7).

Microbiologically, bacterial products secreted from periodontal pathogens could trigger the inflammatory cascade seen in RA, as part of a systemic inflammatory state, as well as the production and spreading of inflammatory mediators by affected periodontal tissues(8). Therefore, periodontal treatment could lead to a decrease in systemic inflammatory mediators, and as a consequence, lower the symptoms of RA.

An association was found, in systematic reviews of case control studies, for inflammatory markers in patients with PD and RA,(9–12). A systematic review assessing the effect of PD treatment on RA outcomes included 7 controlled studies and 1 clinical trial(13), a reduction of DAS28 was observed after PD treatment, this reduction was replicated in a review that found 4 interventional studies, out of which only 1 was a randomised clinical trial(14).

The currently available evidence on this subject is inconclusive. The aim of this systematic review is to provide a rigorous up to date summary of the available evidence on the role of periodontal therapy in the treatment of patients with rheumatoid arthritis.

## Methods

### Protocol and Registration

This manuscript complies with the Preferred Reporting Items for Systematic review and Meta-Analysis for reporting systematic reviews and meta-analyses.

This protocol was adapted to the specificities of the question assessed in this review and registered to PROSPERO with the ID CRD42020150286.

### Elegibility Criteria

#### Types of studies

We will include randomized controlled trials (RCTs) meeting our inclusion criteria and reporting useable data. Quasi-randomized trials (QRTs) will not be included. We will exclude studies evaluating the effects on animal models or in vitro conditions.

#### Types of participants

We will include trials assessing participants aged 18 and above, with a diagnosis of active RA as defined by the American College of Rheumatology 1987 criteria or the 2010 American College of Rheumatology/ European League against Rheumatism classification criteria for rheumatoid arthritis and chronic periodontitis with probing depth ≥ 4mm and clinical attachment loss ≥ 2mm in at least two teeth.

Individuals with necrotising periodontitis and other gingival and periodontal lesions as manifestation of systemic diseases will be excluded.

#### Types of interventions

We will include trials evaluating rheumatoid arthritis outcomes after periodontal treatment comprised of:

- Mechanical debridement (MD): including root surface instrumentation, scaling and root planning (SRP) and curettage.
- Chemical therapy (CT): systemic antibiotic therapy, local antibiotic therapy and/or antimicrobial rinses.
- Behavioural changes (BC) such as oral hygiene instruction and counselling.

Combinations of the aforementioned with at least one mechanical debridement method. Both active phase therapy and maintenance therapy will be included.

The comparison of interest will be placebo/sham (PT with MD versus sham MD) (PT with MD versus PT without MD) or no treatment (PT with MD versus no PT).

Trials assessing periodontal therapy plus other interventions will be eligible if the cointerventions are identical in both intervention and comparison groups. Trials evaluating periodontal therapy in combination with other active interventions versus placebo or no treatment will be also included.

#### Types of outcome measures

We will not use the outcomes as an inclusion criteria, during the selection process. Any article meeting all the criteria except for the outcome criterion will be preliminarily included and assessed in full text.

##### Primary outcomes

- Disease Activity Score 28-joint count (DAS28)

##### Secondary outcomes

- Bleeding on Probing (BOP)
- Clinical Attachment Level (CAL
- Periodontal Probing Depth (PPD)
- Health Assessment Questionnaire (HAQ) or modified versions of the test.

##### Other outcomes

- Total adverse events

#### Search methods for identification of studies

##### Electronic searches

We will conduct a comprehensive search in PubMed/Medline, Embase, Cochrane Central Register of Controlled Trials (CENTRAL), Lilacs, the International Clinical Trials Registry Platform (ICTRP), ClinicalTrials.gov, US National Institutes of Health (NIH) and grey literature, to identify all relevant randomized controlled trials regardless of language or publication status (published, unpublished, in press and in progress). The searches will cover from the inception date of each database until the day before submission.

The following search strategy will be used to search in Pubmed/Medline. We will adapt it to the syntax of other databases.

#1 rheumatoid; #2 RA #3 ((arthritis OR nodul*) AND rheumat*); #4 (felty* AND syndrome); #5 (caplan* AND syndrome); #6 "Arthritis, Rheumatoid"[Mesh]; #7 #1 OR #2 Or #3 OR #4 OR #5 OR #6; #8 periodont*; #9 Periodontitis [MeSH]; #10 scaling; #11 #8 OR #9 OR #10; #12 “randomized controlled trial” [pt]; #13 “controlled clinical trial” [pt]; #14 randomized [tiab]; #15 placebo; [tiab]; #16 “drug therapy” [sh]; #17 randomly [tiab]; #18 trial [tiab]; #19 groups [tiab]; #20 #15 OR #16 OR #17 OR #18 OR #19; #20 (“animals” [mh] NOT “humans” [mh]); #22 #20 NOT #21; #23 #7 AND #11 and #22

##### Searching other resources

An expanded search will be performed to identify articles potentially missed through the database searches and in order to identify ‘grey literature’ and unpublished studies. This includes the following.

- MEDLINE for systematic reviews addressing the same question as our review.
- In order to find additional literature, we will run a Google Scholar search for key terms and authors.
- We will hand search reference lists of all included studies and of relevant reviews retrieved by the electronic searching to identify further relevant trial.
- Authors of included studies will be contacted for any additional published or unpublished data.

#### Selection of studies

The results of the literature search will be uploaded to the screening software Collaboratron^TM^(15).

In Collaboratron^TM^(15), two researchers will independently screen the titles and abstracts yielded by the search against the inclusion criteria. We will obtain full-text reports for all potentially elegible studies that appear to meet the inclusion criteria or require further evaluation to decide about their inclusion. The same review authors will assess independently those articles and decide on fulfilment of inclusion criteria. A third review author will resolve discrepancies in the case of disagreement. Articles retrieved from the screening and included in the review will be recorded in RevMan 5.3(16). Excluded trials after full text revision and the primary reason for the decision will be listed.

The selection process will be documented in a in a PRISMA flow diagram(17) adapted for the purpose of this project.

#### Extraction and management of data

Using standardised forms, two reviewers will independently extract data from each included study. We will collect the following information: study design, setting, baseline characteristics of patients and eligibility criteria; details of intervention, co-interventions and comparison; number of patients assigned to each treatment group, the outcomes assessed and the time they were measured; number of patients with adverse reactions per treatment group and method used to seek adverse reactions. Losses to follow up, exclusions, and the reasons accordingly. A third review author will resolve discrepancies in the case of disagreement.

#### Risk of bias assessment

Risk of bias in included studies will be assessed according to the ‘Risk of bias’ table, which is the tool recommended by The Cochrane Collaboration(18)(19). Descriptions and judgements about the following domains for each study will be included: adequacy of sequence generation, allocation concealment, blinding, addressing of incomplete outcome data, likelihood of selective outcome reporting, and other potential sources of bias (for example if sample size was calculated and if authors reported an intention-to-treat analysis).

The ‘Risk of bias’ table will be prepared independently by two review authors, with a third review author acting as arbiter. Original study authors will be contacted for further information if one of the review authors requires clarification wherever necessary.

#### Measures of treatment effect

Pooled dichotomous outcomes will be reported as risk ratios (RRs) or odds ratios (OR) with 95% confidence intervals (CI). Continuous outcomes will be reported as mean difference (MD) with 95% CI. Outcomes using different scales will be reported as standardised mean differences (SMD) with 95% CI.

Then, these results will be displayed on the ‘Summary of Findings Table’ as mean difference.

#### Dealing with missing data

Main author of trials will be contacted in order to verify key study characteristics and obtain missing numerical outcome data. If not possible, only available data will be analysed. The potential impact of missing data will be addressed in the discussion section, analysis of worst case scenario and best case scenario will be applied.

If no intention-to-treat analysis is reported, or the analysis has been modified, data will be reanalysed following intention to treat principles if possible.

#### Assessment of heterogeneity

Heterogeneity will be quantitatively assessed with a statistical test (Q statistic) and the I^2^ statistic. Statistically significant heterogeneity will be defined as at least one positive test (establishing a cut-off value of P = 0.10 for the Mantel-Haenszel Chi^2^ test, or values over 50% using the I^2^ statistic)

#### Assessment of reporting biases

Publication bias will be evaluated through visual analysis of funnel plots. Evidence of asymmetry will be based on P < 0.10, and present intercepts with 90% CIs. Discrepancies between registered protocols and final publications will be assessed to evaluate other reporting biases including outcome reporting bias. If no record is found for a study in the WHO International Clinical Trials Registry Platform, authors will be contacted for further information.

#### Data synthesis

Statistical analysis will be performed in accordance with the guidelines for statistical analysis developed by Cochrane(19). If applicable, studies will be pooled to perform meta-analysis comparing periodontal treatment versus placebo or no treatment for each outcome measure. When possible, meta-analyses using the random-effects inverse variance model will be carried out to estimate the pooled measure of treatment effect, fixed effects models will be performed to evaluate if findings are not sensitive to choice of analysis. If specific populations or interventions are found to be significantly different, separate meta-analyses will be performed to assess if heterogeneity is explained by some of these, or if a convincing subgroup effect is found. Discrepancies found between random and fixed effects analyses, will be addressed in the discussion.

#### Subgroup analysis and investigation of heterogeneity

If enough data is available, subgroup analysis will be performed to assess any outcome differences depending on initial periodontal disease severity and/or rheumatoid arthritis severity, as well as diabetic and smoking population.

#### Sensitivity analysis

Sensitivity analyses will be performed to evaluate the impact of the inclusion or exclusion of missing data and the choice of a fixed-effect or random-effects models. If there are any substantial differences, data from lower-quality studies will not be pooled with the higher-quality studies, and will be presented in different analyses, unless separate analyses do not differ greatly from pooled results.

#### Summary of Findings

The certainty of the evidence for all outcomes will be reviewed using the Grading of Recommendations Assessment, Development and Evaluation working group methodology (GRADE Working Group)(20). Findings for the main outcomes will be summarised in Summary of Findings (SoF) tables.

## Data Availability

All data related to the project will be available on the final publication.

## Notes

### Roles and contributions

FN conceived the protocol, FN, SL and GC drafted the manuscript, and all other authors contributed to it. The corresponding author is the guarantor and declares that all authors meet authorship criteria and that no other authors meeting the criteria have been omitted.

### Ethics

As researchers will not access information that could lead to the identification of an individual participant, obtaining ethical approval was waived.

### PROSPERO registration

This protocol has been submitted PROSPERO Registration ID: CRD42020150286.

